# Fine-Tuning SAM2 for Coronary Artery Segmentation in X-Ray Fluoroscopy

**DOI:** 10.64898/2026.06.16.26355803

**Authors:** Elakiya Sivakumar

## Abstract

SAM2 [Ravi et al., 2024] (Meta, 2024) provides a strong starting point for segmentation, but given the unique challenges in medical imaging (noise from patient movement, the projection-based nature of X-ray fluoroscopy, and low contrast between vessels and background), direct application is difficult. We fine-tune MedSAM2 [Ma et al., 2025] on annotated coronary angiograms and apply it to video data for point-of-care use. On the ARCADE validation set [Popielarski et al., 2024] (200 images), the fine-tuned model achieves Dice 0.767 ± 0.082 compared to 0.033 zero-shot. In 10 fluoroscopic video studies from CoronaryDominance [Danilov et al., 2025], it tracks vessels coherently and avoids false segmenting of ribs, stents, and bypass grafts in 9 of 10 studies. Code is available [here](https://github.com/elakiyasivakumar/SAM2-Coronary-Angiography-VA) and the fine-tuned checkpoint [here](https://huggingface.co/Elakiya17/CA-SAM2).

## 1 Introduction

Segmentation models have been around since the early 2020s and have been generally used on images and videos, but required one-to-one training for each application. Meta’s 2024 Segment Anything Model [Kirillov et al., 2023] revolutionized the space by being able to segment any object given point click, box prompts, or a combination of both. This model has seen wide utilization across creative and general applications. However, as with most computer vision models, they require fine-tuning for use in specific domains.

Several interventional cardiology procedures require coronary angiography to be performed, with the surgeon re-injecting contrast dye to visualize the vessel path across multiple frames. Computer vision enabled segmentation comes in handy during live procedures to ease physician and technician burden — automated vessel delineation would support real-time guidance and quantitative assessment without manual annotation.

To this effect, this paper attempts to utilize a fine-tuned version of SAM2 and apply it for continuous video imaging applications. We note that there are no publicly available large, well-annotated datasets for interventional cardiology procedures with frame-level ground truth. For the purposes of this paper we use a static angiogram dataset for finetuning and a video dataset for qualitative evaluation.

## 2 Related Work

### Coronary artery segmentation

Classical approaches relied on handcrafted vesselness filters [Frangi et al., 1998] and graph-based tracking. CNN-based methods improved performance but depend on large annotated corpora that are scarce in interventional imaging. The ARCADE challenge [Popielarski et al., 2024] is the main public benchmark for static coronary angiograms. Frame-level video segmentation of fluoroscopy has no established benchmark.

### Foundation models for medical segmentation

SAM [Kirillov et al., 2023] and MedSAM [Ma et al., 2024] demonstrated interactive segmentation on medical images. SAM2 [Ravi et al., 2024] extended this to video via a streaming memory module. MedSAM2 [Ma et al., 2025] adapts SAM2 to medical imaging across CT, MRI, ultrasound, and endoscopy. SurgiSAM2 [Yue et al., 2025] and FS-MedSAM2 [Zhao et al., 2024] show fine-tuning on surgical and general medical video. To our knowledge, X-ray fluoroscopy has not been evaluated in prior work.

### Topology-preserving losses

clDice [Shit et al., 2021] measures centerline overlap via soft skeletonization of predicted and ground-truth masks, preserving the connectivity of thin tubular structures. It has been applied to retinal vessel segmentation [Wang et al., 2022, Iqbal et al., 2024]; its application to coronary angiography is novel.

## 3 Methods

### 3.1 Datasets

**ARCADE [Popielarskietal., 2024].** 1,200 X-ray coronary angiogram images (512 × 512 px) with polygon annotations across 26 artery segments. We use the official 1,000/200 train/val split. Annotations are characterize into binary masks.

**CoronaryDominance [Danilovetal., 2025].** 1,574 multi-view X-ray fluoroscopy studies (20–70 frames, 512 × 512 px). Labels are study-level dominance classifications only; no per-frame vessel masks are provided. Ten studies are used for qualitative video evaluation.

### 3.2 Model

We use MedSAM2 [Ma et al., 2025], a SAM2 model fine-tuned on broad medical imaging data with weights publicly available on GitHub. The choice of MedSAM2 over vanilla SAM2 as a starting point is motivated by a property specific to transformer architectures: unlike CNNs, vision transformers lack built-in inductive biases (locality, translation equivariance), making their representations more sensitive to the pre-training distribution [Li et al., 2022, Matsoukas et al., 2023]. Starting from a model already exposed to medical image characteristics such as grayscale intensity distributions, anatomical structure, and low contrast-to-noise ratios is a closer initialization to the fluoroscopy domain than one trained on natural images. MedSAM2 uses a Hiera-Tiny backbone [Ryali et al., 2023] (12 trunk blocks, 3 hierarchical stages), an FPN neck, and a two-way transformer mask decoder. For video, a 6-frame FIFO memory module propagates masks without re-prompting.

### 3.3 Fine-Tuning

#### Partial encoder unfreeze

We train the last two Hiera trunk blocks (`blocks[10]`, `blocks[11]`) and the FPN neck which translates to roughly the last 10% of the encoder by parameter count, alongside the mask decoder. All earlier blocks remain frozen. This gives ∼13.5M trainable parameters of 38.9M total (34.7%). We apply discriminative learning rates: 5 × 10^*−*5^ for the decoder, 1 × 10^*−*5^ for the neck, and 5 × 10^*−*6^ for the unfrozen trunk blocks, keeping the backbone from moving too fast relative to the task head.

#### Augmentation

5,000 training images are generated from the 1,000 ARCADE originals via five geometric variants per image: original, horizontal flip, vertical flip, 20^*°*^ rotation, and flip + rotation. Masks receive identical transforms. We restrict augmentations to geometric transforms to preserve the integrity of the vessel masks as intensity-based augmentations may risk misaligning mask boundaries with the augmented image appearance.

#### Training

We train with AdamW, *β* = (0.9, 0.999), and weight decay, which helps regularize the pre-trained backbone on a relatively small fine-tuning set. The learning rate follows a cosine annealing schedule over 40 epochs, preventing oscillation as the model converges. Gradient clipping at *ℓ*_*∞*_ = 0.5 is applied to handle occasional spikes from the clDice skeletonization gradients in early epochs. Training uses FP16 mixed precision with a batch size of 4 on an NVIDIA L4 GPU (23.7 GB VRAM).

#### Prompt

A single centroid click derived from the ground-truth mask at training time, with ± 5 px jitter. At inference, the centroid is auto-computed from the brightest center-weighted pixels of the prompt frame. Matching prompt type between training and inference is important — preliminary experiments with bounding-box training followed by click inference confirmed substantial performance degradation.

### 3.4 Loss

In addition to standard segmentation losses, we incorporate clDice [Shit et al., 2021], a topology-preserving loss designed for thin tubular and network structures. Standard overlap-based losses such as Dice are insensitive to topological errors; a vessel segmented as two disconnected fragments can achieve high pixel-level overlap while being anatomically incorrect and clinically unusable. clDice addresses this by operating on the soft skeleton of both the predicted and ground-truth masks rather than the filled regions, directly penalizing breaks in centerline connectivity. Originally validated on retinal vasculature, brain vessel networks, road networks, and neuronal structures, clDice has since been adopted broadly for tubular structure segmentation. To our knowledge, this is the first application of clDice to coronary X-ray angiography.

The combined loss is:

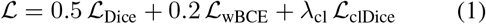

#### Dice (*L*_Dice_)

soft Dice on sigmoid outputs.

#### Weighted BCE (*L*_wBCE_)

BCE with positive-class weigh 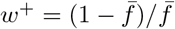, where 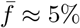 is the mean foreground fraction. Without reweighting, the loss is dominated by background pixels.

#### clDice (*L*_clDice_)

soft skeletonization of predicted and ground-truth masks, penalizing centerline breaks. Computed in FP32 to avoid rounding errors. *λ*_cl_ = 0 for the first three epochs, linearly ramped to 0.3 by epoch 8 — early predictions produce unstable skeletons that destabi-lize training if clDice is applied too soon.

### 3.5 Video Inference

The fine-tuned weights are loaded into MedSAM2’s video predictor, which inherits SAM2’s streaming memory architecture. The prompt frame is selected as the frame with the highest pixel variance, which approximates peak contrast fill. MedSAM2 propagates the mask forward and backward from this frame using its streaming memory.

## 4 Results

### 4.1 Static Segmentation

Table 1 shows Dice and IoU on the 200-image ARCADE validation set. Zero-shot MedSAM2 scores Dice 0.033, which reflects the domain gap between its pre-training modalities and X-ray fluoroscopy. Fine-tuning the decoder alone with a frozen encoder recovers most of the performance at Dice 0.727. Partially unfreezing the encoder and adding clDice loss and augmented training data pushes this to Dice 0.767 ± 0.082 (IoU 0.629). This result is competitive with recent XCA-specific models, which report Dice in the 0.76–0.85 range on comparable coronary segmentation tasks [Zhang et al., 2025, Chang et al., 2024, Kaba et al., 2023], achieved here with a starting model that had never seen X-ray fluoroscopy.

**Table 1:**
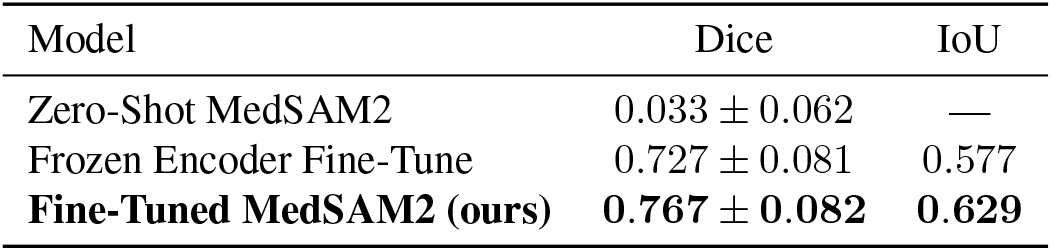
ARCADE validation set results (200 images, centroid click).

### 4.2 Video Inference

Figure 1 shows a representative frame sequence from a CoronaryDominance fluoroscopy study. The full comparison video is available in the code repository.

**Figure 1.**
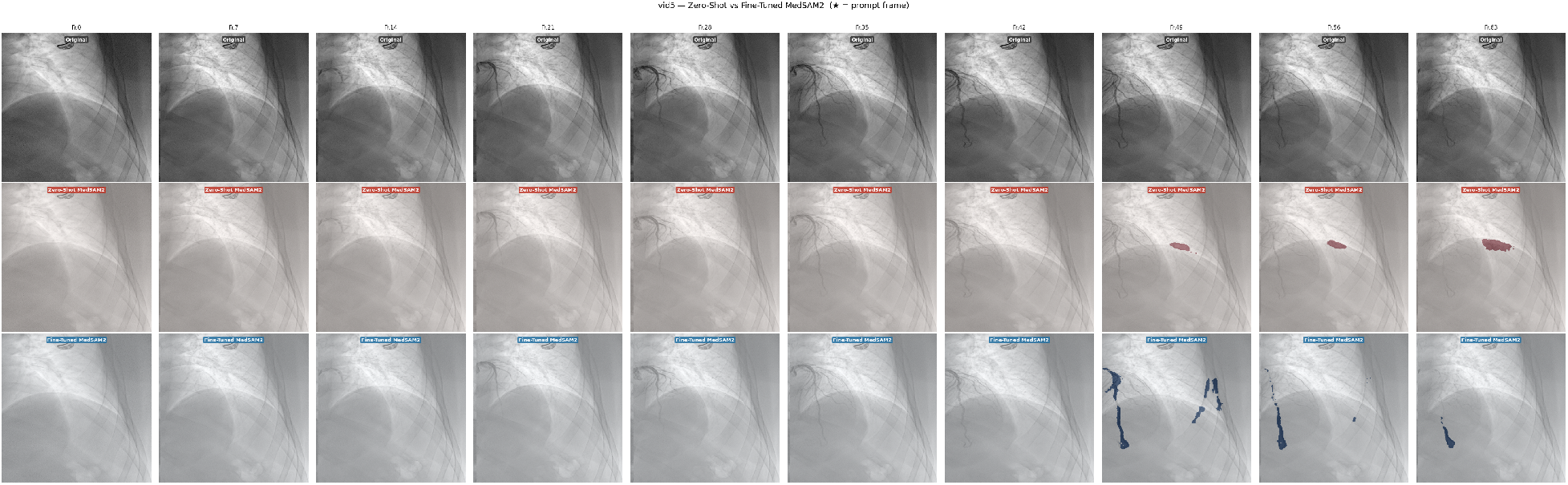
Frame comparison on a representative CoronaryDominance study. *Top*: original fluoroscopy frames. *Middle*: zero-shot MedSAM2, activates on ribs and non-vessel structures. *Bottom*: fine-tuned MedSAM2, vessels tracked with tighter boundaries, false positives absent.

The fine-tuned model tracks the main coronary trunk across frames without re-prompting, with tighter boundary delineation than the zero-shot model. Both models fail when contrast clears between injection cycles. SAM2’s 6-frame memory loses the vessel target once contrast disappears; re-prompting at the start of each injection cycle is the practical fix.

The most striking finding across the studies is specificity. In 9 of 10 studies, the fine-tuned model does not activate on ribs, surgical stents, bypass grafts, or implanted devices — structures the zero-shot model consistently mislabels because their tubular, high-contrast appearance is visually similar to vessels for a model without fluoroscopy-specific training. Fine-tuning on real coronary vessel masks appears to teach the model what vessels actually look like in this modality, rather than what vessel-shaped objects look like in general. Figure 2 shows this directly on a single representative frame.

**Figure 2.**
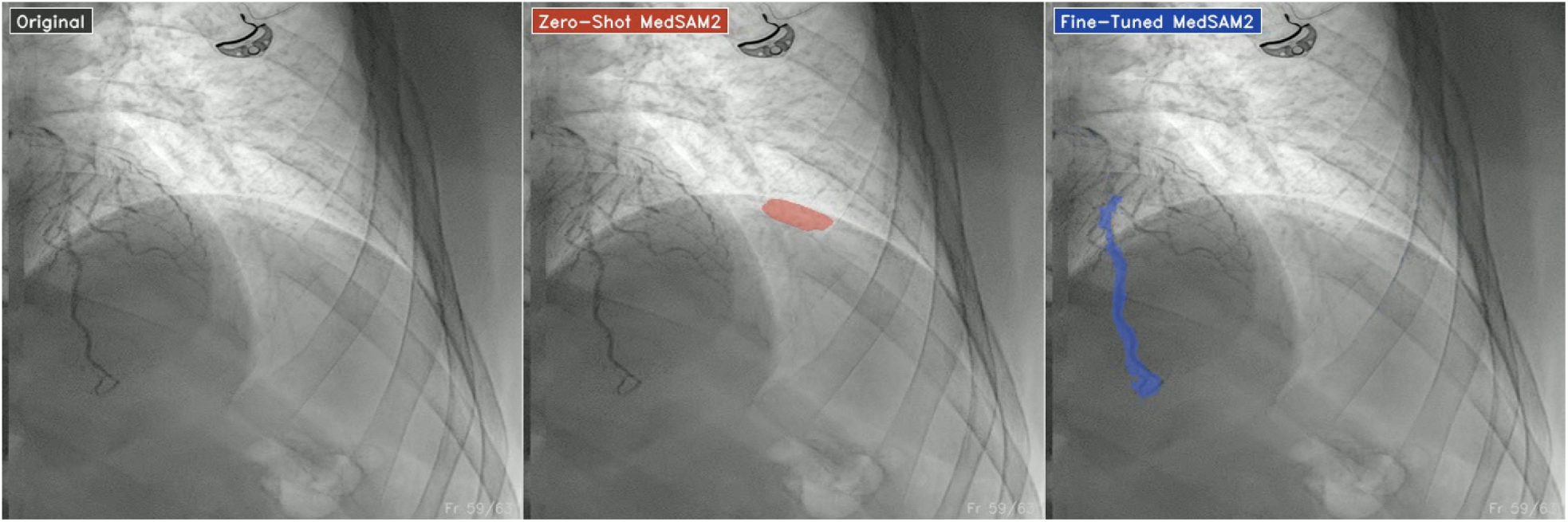
Single-frame comparison on a representative CoronaryDominance fluoroscopy study. *Left*: original fluoroscopy frame. *Center*: Zero-Shot MedSAM2 (red overlay): the model incorrectly activates on an implanted cardiac device rather than the coronary vessel. *Right*: Fine-Tuned MedSAM2 (blue overlay): the model correctly localizes and delineates the coronary vessel with tight boundaries. This failure mode of the zero-shot model was observed in 9 of 10 evaluated studies and is absent in the fine-tuned model.

## 5 Discussion

The gap from Dice 0.033 to 0.767 with only 1,000 training images (5,000 post-augmentation) indicates that Med-SAM2 can adapt to X-ray fluoroscopy without extensive data. The backbone already captures useful low-level structure; partially unfreezing the deepest two blocks is enough to shift high-level representations toward the new modality without overwriting general features.

The specificity result on video is arguably more important than the Dice number. A model that segments stents and ribs as vessels is not usable in a clinical setting regardless of its benchmark score. The observation that fine-tuning on coronary vessel masks suppresses this failure mode — without any explicit negative supervision — suggests the model is learning genuine vessel appearance rather than generalized tubular-object detection.

### Limitations

Currently, our video evaluation results are qualitative and lack numerical metrics such as Dice scores; this is due to CoronaryDominance having no perframe ground truth. Quantitative video evaluation requires clinician-annotated fluoroscopic sequences that are not currently publicly available.

SAM2’s memory-based propagation architecture introduces a tracking latency at the start of each contrast injection cycle. Because the model initializes tracking only after a prompt frame is placed at peak contrast fill, there is a window of milliseconds to seconds — depending on injection rate and frame rate — during which the vessel cannot be reliably located. Addressing this would require either predictive prompting mechanisms or memory architectures that can initialize from partial contrast fill.

Fine-tuning on static images does not directly improve the temporal memory mechanism. The centroid click captures the main trunk but misses the thin distal branches.

### Future work

This work uses supervised pre-training (MedSAM2) as the initialization strategy. A natural next direction is self-supervised pre-training directly on unlabeled fluoroscopic video, large quantities of which exist in clinical archives, to build representations specific to this modality before any supervised fine-tuning. Contrastive or masked-autoencoder approaches on fluoroscopy data could reduce dependence on annotated datasets and potentially push Dice scores beyond what supervised pretraining allows. Video-aware fine-tuning on actual fluoro-scopic sequences, rather than static angiograms, is a further direction for improving temporal coherence.

For point-of-care deployment, the model needs to run on clinical hardware rather than data center GPUs. We hope to explore implementations of smaller SAM variants such as MobileSAM or EfficientSAM for deployment on resource-constrained hardware.

## 6 Conclusion

We fine-tune MedSAM2 on coronary angiograms and apply it to fluoroscopic video. Partial encoder unfreeze, clDice loss, and offline augmentation bring Dice from 0.033 zero-shot to 0.767 on ARCADE. On fluoroscopic video, the model tracks vessels coherently and, in 9 of 10 studies, avoids the false positives on non-vessel structures that are common in the zero-shot model.

## Data Availability

All data produced are publicly available online at ARCADE Popescu, D. et al. ARCADE: Automatic Region-based Coronary Artery Disease diagnostics using X-ray angiography images. Scientific Data 11, 242 (2024). Data: https://zenodo.org/records/10390295 CoronaryDominance Zhu, Z. et al. Coronary Dominance Classification from X-ray Angiography. Kaggle Dataset (2022). https://www.kaggle.com/datasets/zhifanl/coronary-dominance

https://www.kaggle.com/datasets/zhifanl/coronary-dominance

## Acknowledgements

Datasets: ARCADE [Popielarski et al., 2024] (Zenodo 10390295), CoronaryDominance [Danilov et al., 2025].

